# Caudate Volume is Prospectively Associated with Irritability in Toddlerhood

**DOI:** 10.1101/2023.06.17.23291514

**Authors:** Alexander J. Dufford, Leigha MacNeill, Ashley Nielsen, Christopher Smyser, Joan L. Luby, Cynthia E. Rogers, Elizabeth Norton, Lauren Wakschlag

## Abstract

Irritability refers to the dispositional tendency toward angry emotion with both mood and behavioral elements. The dimensional spectrum of irritability is an RDoC-informed transdiagnostic marker of psychopathology risk, specifically the common and modifiable internalizing and externalizing disorders. Despite substantial interest in this robust developmentally based transdiagnostic indicator of psychopathology risk, its early brain markers are understudied. Here, we present data (n=31) from an imaging sub-study of the When to Worry study, in which we examined prospective associations between volume in three subcortical regions implicated in irritability (the caudate, putamen, and amygdala) around the infants’ first birthday (Baseline) and the dimensional spectrum of observed irritability using the Disruptive Behavior Diagnostic Observation Schedule (DB-DOS) around toddlers’ second birthday (Follow-up). Both left (*q*<.04, FDR corrected) and right caudate volumes (*q*<.04, FDR corrected) at Baseline were negatively associated with a measure of irritability measured at Follow-up. We did not find support for associations between putamen and amygdala volumes at Baseline and observed irritability at Follow-up. These findings identify early prospective neuroanatomical correlates of toddler irritability and provide preliminary support for the caudate being an important brain region for understanding the developmental sequalae of irritability.

## Introduction

Irritability is a salient and measurable transdiagnostic marker of later internalizing and externalizing disorders that can be measured reliably early in development (Finlay-Jones et al., 2023; Klein et al., 2021; Nielsen et al., 2021; Wakschlag et al., 2019). Irritability refers to a dispositional tendency towards anger with both mood and behavioral elements (Finlay-Jones et al., 2023; Nielsen et al., 2021; Wakschlag et al., 2018; Wakschlag et al., 2019; Wiggins et al., 2021). Although its most salient feature, tantrums, are a normative misbehavior of early childhood, it is atypical when frequent, disproportionate to context, and dysregulated (Wakschlag, Choi, et al., 2012). Due to its transdiagnostic nature and ability to be measured on a dimensional normal:abnormal spectrum, recent studies have approached studying irritability using the Research Domain Criteria framework (RDoC) (Brotman et al., 2017; Damme et al., 2022; Grabell et al., 2018; Mittal & Wakschlag, 2017; Nielsen et al., 2021; Wakschlag et al., 2015). In particular, to advance with RDoC’s goal of understanding psychopathology across a normal:abnormal spectrum in terms of mechanistic pathways, recent studies have focused on placing irritability as a transdiagnostic indicator of psychopathology within a neurodevelopmental context and examining its neurobiological correlates (MacNeill et al., 2021; Nielsen et al., 2021). While studies have identified both structural and functional correlates of irritability, these studies have primarily focused on irritability in childhood and adolescence (Lee et al., 2022; Perlman et al., 2015). However, the increasing evidence that atypical irritability can be identified behaviorally in the first years of life, aligning with the RDoC framework, now enables interrogation of corollary neural antecedents of early irritability (Finlay-Jones et al., 2023; Krogh-Jespersen et al., 2022; Liu et al., 2018). These neural antecedents prospectively associated with irritability to focus investigations exploring these regions as underlying potential underlying mechanisms or ‘neural markers’ that may be targeted for early therapeutic interventions and perhaps identifiable even earlier than behavioral indicators (Damme et al., 2022; Deveney et al., 2019; MacNeill et al., 2021; Mittal & Wakschlag, 2017).

As mentioned, the examination of the neural correlates of irritability have primarily focused on child and adolescent samples (Lee et al., 2022; Perlman et al., 2015). A systematic review and meta-analysis of the literature examining associations between irritability and brain imaging found the most consistent findings for aberrant functional activation for the amygdala, caudate, and putamen (Lee et al., 2022). Examining l1 studies focusing on structural MRI, the study found little consensus in terms of regions. While these studies have found converging neural correlates in terms of functional correlates of irritability, little is known about structural neural correlates, especially before early childhood. Of the existing studies, correlations between brain structure and irritability have been primarily found in fronto-striatal regions (Dennis et al., 2019; Perlman et al., 2015; Sammallahti et al., 2023). A study focused on the structural neural correlates of irritability found a negative association between irritability (as measured by excessive crying) in infancy and amygdala volume at 10 years old (Sammallahti et al., 2023). Further, a study in 151 adolescents (mean age=11.5) found both negative and positive associations between brain structure and irritability measured with the Affective Reactivity Index (Dennis et al., 2019). The study found negative associations between irritability and gray matter in the putamen, further, in regions that are typically expanding in terms of gray matter during this developmental period, the study found associations between irritability and less expansion in the caudate and insula. In the present study, we aim to examine the early neural structural correlates of irritability by testing prospective associations between volumes of the caudate, putamen, and amygdala at around the child’s first birthday (Baseline, mean age=14.7 months, SD=1.3 months) and clinically salient irritability via a validated observational assessment, the Disruptive Behavior Diagnostic Observation Schedule (DB-DOS) around the child’s second birthday (Follow-up, mean age=26.9 months, SD=2.7 months). Our analysis focused on these three subcortical regions (caudate, putamen, and amygdala) for they are involved in emotion regulation and reward learning processes (Grahn et al., 2008; Haruno & Kawato, 2006) and have previously been identified as neural correlates of irritability in children and adolescents (Lee et al., 2022). Due to the neurodevelopmental trajectories of these subcortical regions across infancy and into toddlerhood (Knickmeyer et al., 2008) and previous studies showing negative brain structure and irritability correlations (Dennis et al., 2019; Sammallahti et al., 2023), we hypothesized lower caudate, putamen, and amygdala volumes at Baseline would be prospectively associated with greater irritability at Follow-up.

## Materials and Methods

### Participants

The current study focused on an MRI sub-sample of participants drawn from a larger longitudinal study of early irritability, “When to Worry (W2W)”. The W2W was comprised of 356 diverse families with a child aged between 12–18 months at Baseline, living in a large urban area of the US Midwest region (Krogh-Jespersen et al., 2022). The sample was enriched for irritability using the Multidimensional Assessment Profile Temper Loss Scale-Infant/Toddler Version (MAP-DB) (Wakschlag, Choi, et al., 2012; Wakschlag, Henry, et al., 2012; Wakschlag et al., 2010). In the full sample, 243 children had usable behavioral data for coding irritability. From the full sample, 204 families were approached about participating in the Baseline session to obtain an MRI scan. The MRI sub-sample was chosen based upon high and low scores of irritability from the larger study based upon scores from the MAP-DB such that recruitment focused on having a representative sample across the irritability spectrum (a recruitment goal of including equal numbers of infants above the 85^th^ percentile and below the 50^th^ percentile for MAP-DB irritability scores. Of the 101 families that came to the MRI scan, imaging data was collected for 48 participants. Of the 48 participants with usable structural MRI data, 13 participants were removed because they did not have DB-DOS data at Follow-up (n=35). Based upon the initial visual quality control, two participants were removed for having insufficient quality for analysis due to motion artifact (n=33). From the sample that passed visual quality control, one participant was removed due to poor segmentation quality, and one participant was removed as they did not have any income information collected. This resulted in the final analytic sample of n=31. Demographic information for the analytic sample (usable structural MRI data and DB-DOS) is presented in **Table 1**. We compared the final analytic sample (n=31) to the full W2W sample not included in the imaging portion (n=379) for participant sex (MRI final analytic sample=58.1% male, full W2W sample=54.1% male), the Chi-square test was nonsignificant (*p*<.64). We used Fisher’s exact tests to test for participant race (MRI final analytic sample=45.2% White/Caucasian, full W2W sample=52.7% White/Caucasian) and participant ethnicity (MRI final analytic sample=38.7% Hispanic, full W2W sample=23.7% White/Caucasian) representativeness of the final analytic sample. The exact tests were nonsignificant for race (*p*<.09) and significant for ethnicity (*p*<.04). This indicated that the final analytic sample included more participants with Hispanic ethnicity. For the continuous variables, t-tests with Satterthwaite correction were significant for income-to-needs ratio (*p*<.008) with a mean of MRI final analytic sample=3.1 and mean of full W2W sample=4.4, indicated slightly lower income-to-needs ratio in the final analytic sample. Participants in the final analytic sample did not differ from the full sample for age at irritability assessment (*p*<0.87) with a mean of 26.95 for the final analytic sample and 26.88 for the full W2W sample. Lastly, the final analytic sample did not significantly differ from the full W2w in terms of observed irritability as measured by the Anger Modulation factor score on the DB-DOS (*p*<.70), with a mean of -0.25 for the final analytic sample and a mean of -0.33 for the full W2W sample. Approval from Northwestern University’s Institutional Review Board was received prior to the start of this study (IRB# STU00202880).

**Table 1.**
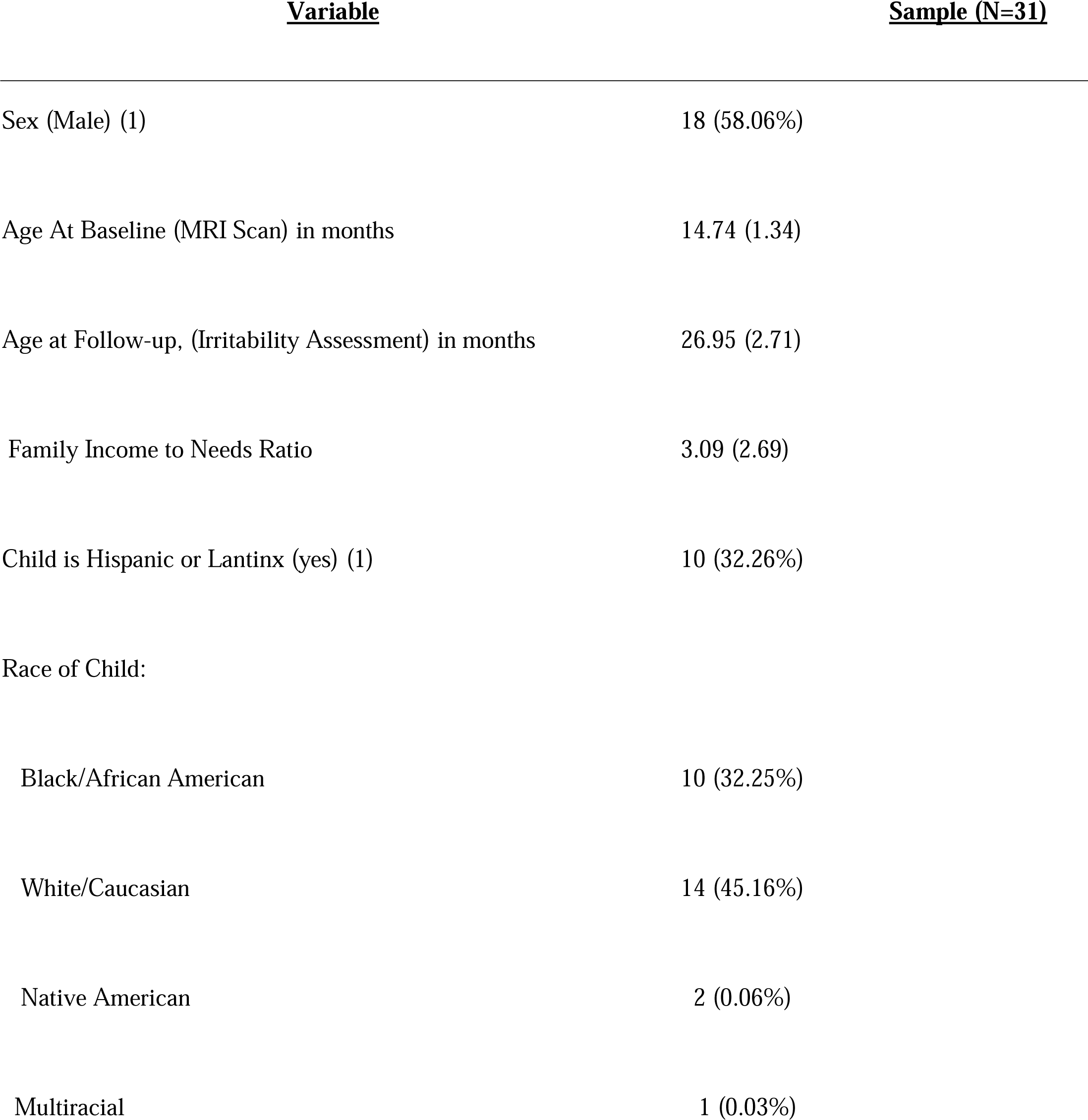
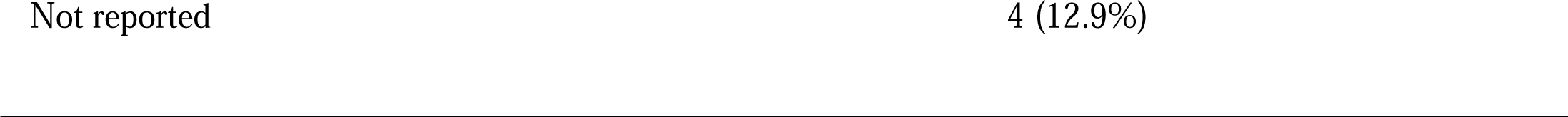
Demographics of the When to Worry neuroimaging final analytic sample.

| <u>Variable</u> | <u>Sample (N=31)</u> |
| --- | --- |
| Sex (Male) (1) | 18 (58.06%) |
| Age At Baseline (MRI Scan) in months | 14.74 (1.34) |
| Age at Follow-up, (Irritability Assessment) in months | 26.95 (2.71) |
| Family Income to Needs Ratio | 3.09 (2.69) |
| Child is Hispanic or Lantinx (yes) (1) | 10 (32.26%) |
| Race of Child: |  |
| Black/African American | 10 (32.25%) |
| White/Caucasian | 14 (45.16%) |
| Native American | 2 (0.06%) |
| Multiracial | 1 (0.03%) |

### Disruptive Behavior Diagnostic Observation Schedule (DB-DOS)

To assess observed toddler irritability, mothers and children completed the well-validated DB-DOS (Wakschlag, Briggs-Gowan, et al., 2008; Wakschlag, Hill, et al., 2008). The DB-DOS is standardized clinical observation specifically designed to characterize observed patterns of behavioral and emotional (dys)regulation that distinguish typical and atypical features across interactional and motivational contexts (Petitclerc et al., 2015). The DB-DOS was originally designed to elicit, observe, and assess disruptive behavior in preschoolers. Adapted versions of the DB-DOS have since been developed to capture observed irritability as transdiagnostic risk pattern and for younger ages including the toddler version used here. Dyads engaged in several tasks that vary in motivational salience, including puzzles, bubble play, “busy mom” (i.e., waiting for parent attention), finger painting, clean-up, and free play tasks.

Child irritability was coded using The Problems in Anger Modulation coding system, capturing behaviors reflecting modulation of irritable affect and behavior from normative variation to clinically concerning). This system incorporates 6 codes: Intensity of Irritable/Angry Behavior (highest level of behavior observed, e.g., uncontrollable crying, throwing objects etc.); Predominance of Irritable/Angry Behavior (e.g., the pervasiveness of irritable/angry behavior across the context); Ease of Elicitation of Irritable/Angry Behavior (both the frequency and precipitant of behavior); Rapid Escalation of Irritable/Angry Behavior (the rate at which irritability escalates); Difficulty Recovering from Irritable/Angry Behavior (how quickly and easily a child recovers); and Copes with Frustration Poorly (how the child manages frustration, e.g., coping strategies such as self-soothing behavior like thumb sucking). As this code was originally developed to assess irritability in preschoolers, the anchor points for some of the existing codes were adapted for the 24-month version to make the codes more developmentally appropriate. For example, tantrums, throwing or banging toys, and sustained/intense crying replaced some of the behaviors included in the preschool manual such as angry tone vocalizations, sighing, and sullenness. Behaviors are rated across an ordinal scale: 0 (None, or normative variation), 1 (Low, or normative misbehavior), 2 (Moderate, or of concern), 3 (High, or clearly atypical) (Wakschlag, Briggs-Gowan, et al., 2008; Wakschlag, Hill, et al., 2008).

Trained coders, led by a master coder, coded these data. Inter-rater reliability was completed on 19% of the IDs. The linear weighted kappa statistics were as follows: Intensity of Irritable/Angry Behavior (0.73), Predominance of Irritable/Angry Behavior (1.00), Ease of Elicitation of Irritable/Angry Behavior (0.94), Rapid Escalation of Irritable/Angry Behavior (0.83), Difficulty Recovering from Irritable/Angry Behavior (0.97), and Copes with Frustration Poorly (0.92). The irritability factor score at 24 months was scored using the calibrations from the 12-month time point. Specifically, we fitted a model in which all the parameters for the 2-year-old model were fixed to the estimates obtained from the 1-year-old model. The 1-year-old model employed a confirmatory factor analytic framework with robust maximum likelihood estimation using Mplus version 8.7. The six codes, modeled as ordinal indicators, loaded onto the primary factor, while Intensity and Rapid Escalation of Irritable/Angry Behavior also loaded equally onto a second uncorrelated factor. This second factor was included to account for the high relationship between the two codes.

### MRI Procedure, Acquisition, and Quality Control

Infants were scanned at the Northwestern University Feinberg School of Medicine’s Center for Translational Imaging. Imaging was conducted during their natural sleep, without sedation, using the “feed and swaddle” technique (Antonov et al., 2017; Korom et al., 2022; Spann et al., 2022). T1-weighted structural images (MP-RAGE) were acquired on a 3T Siemens Prisma^fit^ with a 32-channel head coil with the following parameters: 0.8 mm isotropic resolution, TR/TE = 2400/3.19 ms, and flip angle = 8°. T1-weighted images were repeated if they were determined to not be of sufficient quality and if the toddler remained asleep. Each T1-weighted image was visually inspected using a previously validated quality rating system (Blumenthal et al., 2002).

### Structural Image Analysis

T1-weighted images that passed the visual inspection quality control procedure were processed using an infant/toddler-specific version of Freesurfer (Zöllei et al., 2020). The technical details of the pipeline have been reported previously (Zöllei et al., 2020). Briefly, infantFS uses the T1-weighted image for volumetric segmentation. T1-weighted images are skull-stripped using a supervised skull-stripping algorithm based upon three convolutional neural networks. Volumetric segmentation is performed using a multi-atlas label fusion segmentation framework. The volumetric segmentation concludes with quantifying the volume of multiple subcortical regions in alignment with the standard Freesurfer pipeline’s ‘aseg’ output. We used the absolute volume of the structures to aid in interpretability but included total intracranial volume as a covariate to control for global effects. The infantFS pipeline has been previously shown to be appropriate for data collected between 0 and 2 years of age (Zöllei et al., 2020). One of the authors (AJD) performed quality control for reach segmentation by examining each segmentation slice-by-slice and determining outliers by removing any participants with amygdala, caudate, or putamen volumes that were greater than 2 standard deviations from the mean.

### Statistical Analyses

We examined associations between demographics variables (toddler’s age at scan, age at the administration of the DB-DOS, sex, income-to-needs ratio, and total intracranial volume), subcortical volumes, and irritability using Pearson correlations. We used six multiple regressions (3 regions of interest, left and right for each region) to test for prospective associations between subcortical volumes at Baseline (caudate, putamen, and amygdala) and observed irritability at Follow-up. Each regression included the toddler’s age at scan, age at the administration of the DB-DOS, sex, income-to-needs ratio, and total intracranial volume. To correct for multiple comparisons, the *p*-values from the six multiple regression models were adjusted to control the False Discovery Rate (FDR) at 0.05 using the Benjamini-Hochberg procedure (Benjamini & Hochberg, 1995).

## Results

We examined the associations between the demographic and outcome variables (DB-DOS) using Pearson correlations. All associations were not significant (*p*s>0.05) except for a significant positive association between age at DB-DOS assessment and DB-DOS irritability scores *r*(29)=-.39, *p*=.02. The multiple regressions testing the association between left and right putamen volume (at Baseline) and irritability (at Follow-up were nonsignificant (*p*=.61 and *p*=.25, respectively). The association between left and right amygdala volume (at Baseline) and irritability (at Follow-up) were also nonsignificant (*p*=.59, and *p*=.27, respectively). The multiple regression testing of the left caudate volume’s association with irritability was significant (*R^2^*=.44, *F*(6,24)=3.26, *p*=.01, see **Figure 1**). This indicated a significant prospective negative association between left caudate volume and irritability (*b*=-1.42, *t*(30)=-2.65, *p*=.01). Further, the regression testing the right caudate volume’s association with irritability was significant (*R^2^*=.46, *F*(6,24)=3.50, *p*=.01). Right caudate volume was significantly associated with irritability (*b*=-1.53, *t*(30)=-2.80, *p*=.00). After FDR correction, the *q* values for both the left and right caudate’s association with irritability were significant (*q*<.04 and *q*<.04, respectively).

**Figure 1.**
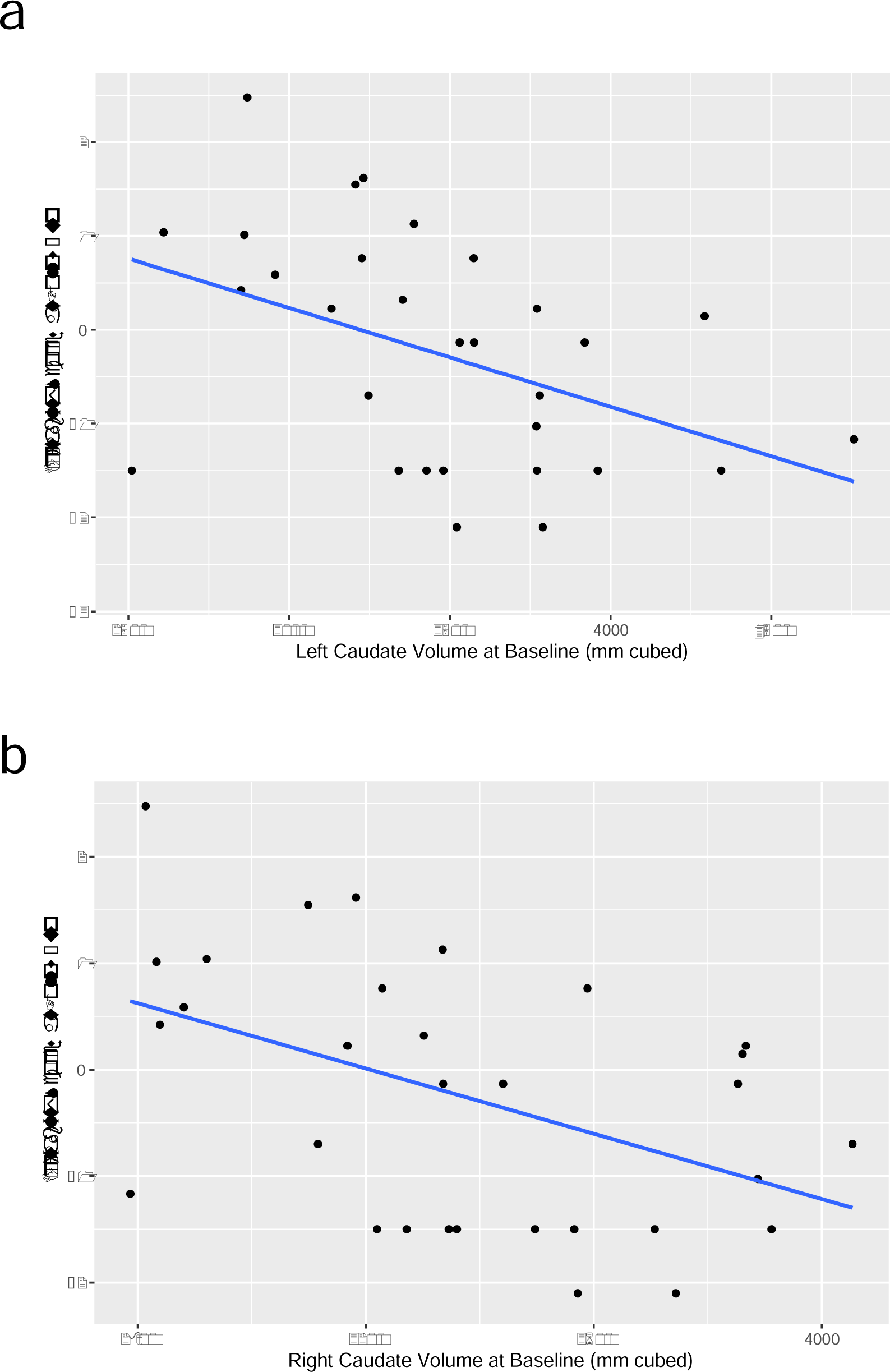
(a) Scatter plot of the prospective negative association between left caudate volume at age 1 and irritability at Follow-up. (b) Scatter plot of the prospective negative association between right caudate volume at Baseline and irritability at Follow-up.

## Discussion

As irritability has been shown to be an early transdiagnostic indicator of psychopathology, it is critical to understand its neural correlates, especially those that may be prospectively associated. We found evidence that both left and right caudate volume at Baseline (around the child’s first birthday) has a prospective and negative association with irritability, at Follow-up (around the child’s second birthday). We did not find support for prospective associations between two other subcortical structures implicated in irritability, the putamen and amygdala, and irritability at Follow-up. These findings identify the volume of the caudate as a prospective neural correlate of irritability; critically this relation is evident in toddlerhood, suggesting its potential for future investigations regarding the early neural mechanisms of irritability.

The goal of the study was to begin to address the gap in understanding the neural correlates of irritability by examining associations before early childhood. Our findings of a prospective negative association between caudate volume and irritability expand upon previous studies in children and adults that have also identified the caudate as a neuroanatomical correlate of irritability (Dennis et al., 2019; Sammallahti et al., 2023), consistently find a negative association, and demonstrate its detection in infancy. While the current study only examined brain structure, the function of the caudate has been consistently shown in childhood and adolescence to have aberrant functional activation in reward processing tasks (Lee et al., 2022). For example, the caudate has been hypothesized to play a role in the difficulties with reward processing involving fronto-striatal regions including the prefrontal cortex, cingulate gyrus, and putamen. Studies of brain function in clinical populations have also reported decreased striatal activation during frustration tasks (Deveney et al., 2019; Perlman et al., 2015). These findings are thought to reflect irritability as a mapping on to the negative valence construct of in the RDoC matrix known as “frustrative nonreward”. These studies, in combination with our evidence of the caudate as an early neural correlate of irritability, motivate future studies to examine associations between the functional connectivity and activation of the caudate and irritability into toddlerhood and childhood.

In the first two years of life, subcortical regions of the brain are undergoing rapid and unparalleled development (Knickmeyer et al., 2008). The caudate’s volume increases by 19% from the first year to second year of life (Knickmeyer et al., 2008). Thus, the results from the current study suggest that toddlers with larger caudate volume have lower levels of irritability. A study of adolescents, scanned longitudinally around ages 11 and 13, found that higher levels of irritability were associated with less expansion and more contraction of the caudate (Dennis et al., 2019). This aligns with the findings of the current study providing convergent evidence that greater volume of the caudate is associated with lower levels of irritability. However, the difference in developmental stage adds additional complexity as subcortical regions in infancy are typically expanding while regions in this period of adolescence are typically decreasing. However, what is consistent is a finding of ‘delayed’ development of the caudate and associations with caudate volume. This aligns with literature showing positive associations between caudate volume and reward processing behaviors such as delay discounting (Tschernegg et al., 2015) and behavioral activation/inhibition (Ide et al., 2020). How the volume, as measured by MRI, of the caudate relates to variations in neuronal structure remains unclear. Using translational models that combine human and animal studies, such as proposed by Brotman et al., 2017, may be used to understand links between irritability, frustrative nonreward, microstructure (neuronal level) and macrostructure (subcortical volume).

A significant strength of the current study was the measurement of irritability using a direct observational assessment. The use of direct observation offers an objective evaluation of nuanced and developmentally sensitive behavioral patterns from typical to atypical that may be less amenable to detection by parent report (Wakschlag et al., 2015). Further, research has shown differential associations of neuroimaging with observed behavior versus parent reports of functioning. For instance, Filippi et al. (2021) found links between amygdala connectivity and observed infant reactivity, but relations with parent-reported temperament were not significant. The DB-DOS captures clinically salient information across a variety of parent-child contexts that vary in the demands placed on both the parent and child. Ratings fall on a continuum of normative variation to clinically concerning behavior, adapted specifically for the toddlerhood period. This finding is aligned with recent calls for ‘deep phenotyping’ of behavior, especially in neuroimaging studies examining brain-behavior associations which have shown to be affected by poor phenotypic reliability and validity (Nikolaidis et al., 2022). The significant association between caudate volume and clinically relevant irritable behavior provides an important developmental context for identifying neural correlates of behavioral risk for emerging psychopathology.

We did not find evidence of prospective associations between putamen or amygdala volumes (at Baseline) and irritability (at Follow-up). As part of the dorsal striatum along with the caudate, the putamen has a differential role in instrumental learning. Evidence suggests that while the caudate integrates information about performance during a task and is involved in the cognitive control demands, the putamen is involved in how likely conditioning stimuli lead to correct responses (Brovelli et al., 2011). Caudate and putamen volumes increase dramatically across the first year of life but the caudate compared to the putamen shows minimal growth in the second year of life (Knickmeyer et al., 2008). Compared to the caudate, aberrant structural volume of the putamen has been primarily linked to ADHD, stereotyped behaviors in ASD, and in preterm birth (Vlasova et al., 2017). Therefore, more in depth, larger, and more representative studies of the differential associations between caudate, putamen, frustrative nonreward, and irritability are needed. In proposed models of aberrant neurobiological functioning in irritability, evidence suggests that in addition to differences in reward processing, youth with irritability also have disrupted threat processing (Brotman et al., 2017). Neuroimaging studies in children and adolescents have identified the amygdala and hippocampus to be associated with irritability both in terms of structural volume (Dennis et al., 2019) and functional activation (Lee et al., 2022). For our study’s findings, we can only speculate as to the absence of associations between putamen/amygdala at Baseline and irritability at Follow-up. One potential contributing factor to the lack of association is that our measure of irritability did not directly measure threat processing which may be more linked to amygdala structure (Brotman et al., 2017; Leibenluft, 2017); however, this may also be due to the limited power due to the small sample size. Further, differences may be methodological, or measurement based as these studies primarily did not use observational measures of irritability as in the present study.

The findings of the current study should be considered considering the following limitations. First, the sample size is modest as obtaining usable neuroimaging data from 1-year-olds remains a challenge (Spann et al., 2022). Second, the mapping between subcortical volume and function is not clear, therefore any inferences in terms of the meaning of the findings for function of the caudate are speculative; therefore, these findings will be tested in future studies using functional MRI. Third, as irritability is an early transdiagnostic indicator of INT/EXT later in development, it will be critical to examine prospective associations between caudate volume, irritability, and internalizing/externalizing symptoms. However, while the W2W study has behavioral data collected at age 3 years, we were not sufficiently powered to examine these associations as only 11 participants had usable MRI data, DB-DOS assessment, and internalizing/externalizing symptoms at age 3 years. Large population-based consortia studies with extensive infant imaging, assessment of irritability, and deep phenotyping such as the HEALthy Brain and Child Development Study (HBCD), will provide outstanding opportunities for replication and extension (Morris et al., 2020).

## Conclusions

Identification of the early neuroanatomical correlates of irritability may provide important insight to underlying neurobiological mechanisms of individual variability in irritability in addition to potentially observe difference in the brain that are precursors to irritability and internalizing/externalizing later in development. These preliminary findings point to exciting new directions for brain:behavior transdiagnostic pathways to psychopathology in early life.

## Data Availability

The data that support the findings of this study are available on request from the corresponding author.

## Acknowledgements

The authors would like to thank the families that participated in the When To Worry study and the funding that supported the study (R01MH107652). We would also like to acknowledge Todd Parrish and Rachael Young for their support in the MRI acquisition at the Center for Translational Imaging – Feinberg School of Medicine. Additionally, we would like to acknowledge Sheila Krogh-Jespersen, James Burns, Yudong Zhang, Amanda Nili, Renee Edwards, Rachel Ahrenholtz, and Erica Anderson.

